# CovidNudge: diagnostic accuracy of a novel lab-free point-of-care diagnostic for SARS-CoV-2

**DOI:** 10.1101/2020.08.13.20174193

**Authors:** Malick M Gibani, Christofer Toumazou, Mohammadreza Sohbati, Rashmita Sahoo, Maria Karvela, Tsz-Kin Hon, Sara De Mateo, Alison Burdett, K Y Felice Leung, Jake Barnett, Arman Orbeladze, Song Luan, Stavros Pournias, Jiayang Sun, Barnaby Flower, Judith Bedzo-Nutakor, Maisarah Amran, Rachael Quinlan, Keira Skolimowska, Robert Klaber, Gary Davies, David Muir, Paul Randell, Derrick Crook, Graham P Taylor, Wendy Barclay, Nabeela Mughal, Luke S P Moore, Katie Jeffery, Graham S Cooke

**Affiliations:** Department of Infectious Disease, Imperial College London, United Kingdom; Imperial College Healthcare NHS Trust, United Kingdom; DnaNudge Ltd, Translation and Innovation Hub, Imperial College White City Campus, London; Department of Electrical and Electronic Engineering, Imperial College London; Chelsea & Westminster NHS Foundation Trust, London; Nuffield Department of Medicine, Oxford University, Oxford, United Kingdom; Oxford University Hospitals NHS Foundation Trust, Oxford, United Kingdom

**Keywords:** SARS-CoV-2, COVID-19, point-of-care diagnostics

## Abstract

3.

**Background:** Access to rapid diagnosis is key to the control and management of SARS-CoV-2. Reverse Transcriptase-Polymerase Chain Reaction (RT-PCR) testing usually requires a centralised laboratory and significant infrastructure. We describe the development and diagnostic accuracy assessment of a novel, rapid point-of-care RT-PCR test, the DnaNudge® platform CovidNudge test, which requires no laboratory handling or sample pre-processing.

**Methods:** Nasopharyngeal swabs are inserted directly into a cartridge which contains all reagents and components required for RT-PCR reactions, including multiple technical replicates of seven SARS-CoV-2 gene targets *(rdrp1, rdrp2, e*-gene, *n*-gene, n1, n2 and n3) and human ribonuclease P (RNaseP) as positive control. Between April and May 2020, swab samples were tested in parallel using the CovidNudge direct-to-cartridge platform and standard laboratory RT-PCR using swabs in viral transport medium. Samples were collected from three groups: self-referred healthcare workers with suspected COVID-19 (Group 1, n=280/386; 73%); patients attending the emergency department with suspected COVID-19 (Group 2, n=15/386; 4%) and hospital inpatient admissions with or without suspected COVID-19 (Group 3, n=91/386; 23%).

**Results:** Of 386 paired samples tested across all groups, 67 tested positive on the CovidNudge platform and 71 with standard laboratory RT-PCR. The sensitivity of the test varied by group (Group 1 93% [84-98%], Group 2 100% [48-100%] and Group 3 100% [29-100%], giving an average sensitivity of 94.4% (95% confidence interval 86-98%) and an overall specificity of 100% (95%CI 99-100%; Group 1 100% [98-100%]; Group 2 100% [69-100%] and Group 3 100% [96-100%]). Point of care testing performance was comparable during a period of high (25%) and low (3%) background prevalence. Amplification of the viral nucleocapsid (n1, n2, n3) targets were most sensitive for detection of SARS-CoV2, with the assay able to detect 1×10^4^ viral particles in a single swab.

**Conclusions:** The CovidNudge platform offers a sensitive, specific and rapid point of care test for the presence of SARS-CoV-2 without laboratory handling or sample pre-processing. The implementation of such a device could be used to enable rapid decisions for clinical care and testing programs.

**RESEARCH IN CONTEXT:** *Evidence before this study:* The WHO has highlighted the development of rapid, point-of-care diagnostics for detection of SARS-CoV-2 as a key priority to tackle COVID-19. The Foundation for Innovative Diagnostics (FIND) has identified over 90 point-of-care, near patient or mobile tests for viral detection of SARS-CoV-2. However, the most widely available rapid tests to date require some sample handling which limits their use at point-of-care. In addition, pressure on supply chains is restricting access to current diagnostics and alternatives are needed urgently.

*Added value of this study:* We describe the development and clinical validation of COVID nudge, a novel point-of-care RT-PCR diagnostic, evaluated during the first wave of the SARS-CoV-2 epidemic. The platform is able to achieve high analytic sensitivity and specificity from dry swabs within a self-contained cartridge. The lack of downstream sample handling makes it suitable for use in a range of clinical settings, without need for a laboratory or specialized operator. Multiplexed assays within the cartridge allow inclusion of a positive human control, which reduces the false negative testing rate due to insufficient sampling.

*Implication of the available evidence:* Point-of-care testing can relieve pressure on centralized laboratories and increase overall testing capacity, complementing existing approaches. These findings support a role for COVID Nudge as part of strategies to improve access to rapid diagnostics to SARS-CoV-2. Since May 2020, the system has been implemented in UK hospitals and is being rolled out nationwide.

## 5. INTRODUCTION

Since its emergence in December 2019, SARS-CoV-2 has led to over 18,000,00 confirmed cases of COVID-19 and 700,000 deaths by the end of July 2020.^1,2^ Improved access to diagnostics is key to controlling ongoing transmission. The viral load in the upper respiratory tract appears to be highest at – or shortly before – the onset of symptoms^3-5^ and the majority of patients with COVID-19 are diagnosed using reverse transcriptase polymerase chain reaction (RT-PCR) from nasopharyngeal and/or oropharyngeal swabs.

Since the publication of the first genome sequence, several in-house and commercial diagnostic kits have been deployed globally.^6,7^ Laboratory RT-PCR remains the standard of care for detection of SARS-CoV-2, although false negative tests can occur in patients presenting with a clinical syndrome compatible with COVID-19.^8^ However, standard RT-PCR is time-consuming and – where they are available – the technical requirements usually require centralized diagnostic laboratories. Laboratory based tests typically take 4-6 hours to complete, and the transport of clinical samples can mean the turnaround time is frequently over 24 hours,^9^ potentially resulting in delay to diagnosis and inappropriate infection control precautions. An additional limitation to several commercial kits is the lack of a human gene target to control for sample adequacy control (such as Ribonuclease P, *RNaseP)*, thereby failing to identify inadequate samples and contributing to false negative results.^10,11^

Point-of-care (POC) diagnostics can have an impact on patient management and control of infectious disease epidemics^12^ and were identified by a WHO expert group as the first of eight research priorities in response to the COVID-19 outbreak.^13^ POC diagnostics accelerate clinical decision making, enabling effective triage and timely therapeutic and infection control interventions^14^ alleviating pressure on overburdened centralized labs and allowing testing in community settings. However, many existing POC diagnostics still require some sample processing which limit their use.^9,15^

In response to the SARS-CoV-2 pandemic, the CovidNudge® point-of-care platform (DnaNudge Ltd, UK) was redesigned, from its previous commercial use in human DNA typing, to provide true sample-to-answer multiplex RT-PCR diagnosis of SARS-CoV-2, without the need for any laboratory facilities and trained personnel.^16,17^ To assess the performance of this novel diagnostic platform we conducted a diagnostic accuracy study for the diagnosis of SARS-CoV-2 infection against laboratory-based RT-PCR.

## METHODS

### CovidNudge point of care test for SARS-CoV-2

The platform comprises two components: the DnaCartridge and a processing unit (NudgeBox) (**Figure 1**). The DnaCartridge (25×78×85mm; 40g) is a disposable, sealed, and integrated lab-on-chip device that enables sample-to-result PCR. The DnaCartridge consists of two main parts: an amplification unit (AU) and a sample preparation unit (SPU). A swab is immediately inserted directly into the swab chamber of the SPU at the time of collection. The swab is broken leaving the swab tip and the sample within the chamber, which is then sealed. Cartridges are placed in the processing unit (NudgeBox, 28×15.5×13.5cm; 5kg), which provides the pneumatic, thermal, imaging and mechanics required to run an RT-PCR reaction outside of a laboratory setting. The SPU consists of a rotatable mixing unit and circumferentially distanced chambers, containing buffers to extract and purify RNA from the swab sample, as well as a lyophilised PCR master-mix to mix with the extracted RNA (**Figure 1B**). The SPU mixing chamber fits on top of a motor-driven spigot in the NudgeBox, which rotates the mixing unit through each stage of sample processing before filling the wells of the AU, inside which the PCR reaction takes place. Exposed surfaces of the instruments are cleaned regularly between operators with 10% bleach, followed by an isopropyl alcohol wipe to remove any residual bleach. Following the test, the single-use cartridge is disposed of following standard laboratory disposal procedures.

**Figure 1.**
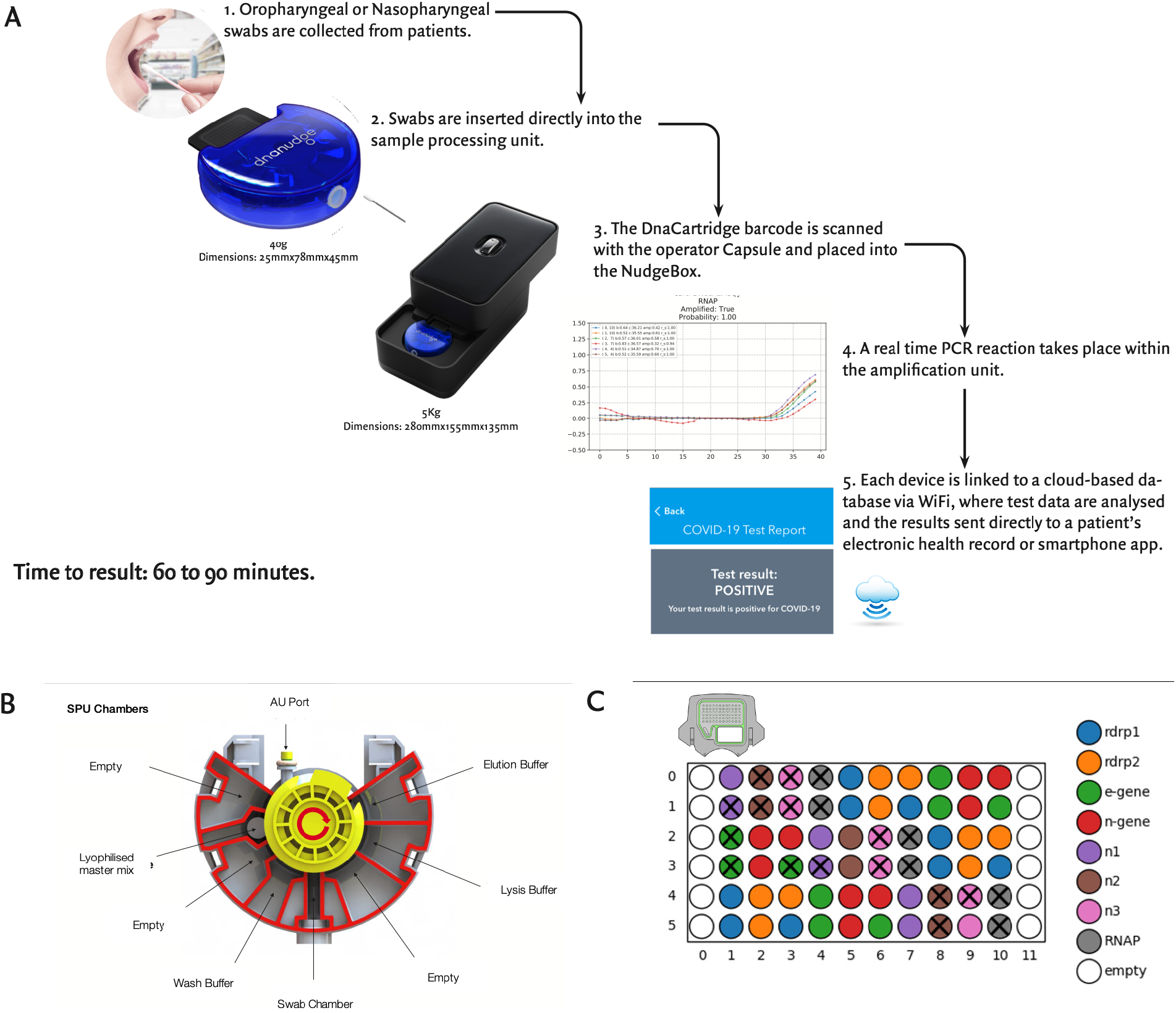
CovidNudge point of care diagnostic for SARS-CoV-2. (A) Schematic of the work-flow. A swab is collected and loaded directly into the DnaCartridge, comprising a sample preparation unit (SPU) and amplification unit (AU). The DnaCartridge is placed into a slot on the lower half of the nudge box, where its SPU mixing chamber fits on top of a motor-driven spigot and the AU sits on top of an active heating and cooling plate. The spigot also connects the DnaCartridge mixing chamber to the pneumatic subsystem. By sliding the upper half to close the NudgeBox, the imaging system aligns on top of the DnaCartridge AU. The upper half also consists of a thermal subsystem which is thermally connected to a mesh plate sitting on top of the AU, which drives the PCR reaction. Data are delivered by WiFi to a cloud-based analysis platform and results are delivered directly to a patients electronic health record. (B) Schematic of sample preparation unit (SPU). The test starts with moving the lysis buffer to the swab chamber. The lysis kills and deactivates the (viral) sample and releases the sample RNA. Silica frit filters are mounted on to the port in the mixing chamber which can capture RNA molecules. The lysis buffer moves from the swab chamber to the mixing chamber and the extracted RNA strands bind to the silica frit filter. In the next step, wash buffer is passed through the mixing chamber and any debris is removed. In the third step, the elution buffer releases the RNA strands from the frit. The elution buffer containing the sample RNA is used to reconstitute the lyophilised RT master mix. In the last step of sample preparation, the mixing chamber turns toward the AU filling port of the SPU to fill the AU. (C) Schematic of the amplification unit (AU). The wells are formed by sealing a mesh membrane to the bottom of the chassis, each less than 1.8uL in volume. Primers and probes for each assay are spotted in nanoliters into the wells, and air dried. To provide redundancy and increase reliability, they are distributed into several wells. The spotting pattern is used by the algorithm to analyse the PCR amplification signals. Each well is represented by a circle coloured according to its assay deposition. Crossed wells indicate target replicated that have amplified in a specific reaction.

The AU comprises dried primers and probes uniquely spotted into 72 reaction wells providing multiplex analysis (**Figure 1C**, **SUPPLEMENTARY METHODS**). For the SARS-CoV-2 assay, the array consists of seven viral targets *(rdrp1, rdrp2, e*-gene, *n*-gene, n1, n2 and n3)^7,18,19^ and one host gene as a positive control assay (Ribonuclease P, *RNaseP)*. Each target has between six to nine technical replicates. The AU sits on top of an active heating and cooling plate, which drives the thermal cycling conditions for the PCR reaction. Multiple cycles of PCR are performed generating florescence data similar to conventional PCR instruments (**Figure 1**).

For a well to be classified as having amplified, the amplification curve should reflect the exponential growth and decay of a standard PCR reaction.^20^ A test is considered valid if ≥3/6 replicates of human RNaseP amplify, reflecting adequate mucosal sampling (see **SUPPLEMENTARY METHODS**). If two or fewer replicates amplify, it is assumed that sample collection was inadequate and the test labelled as invalid. We defined a positive test when ≥2 replicates of at least one viral gene target amplified, otherwise a test was considered negative for SARS-CoV-2.

### Study Design & Participants

Clinical assessment took place at three sites in the United Kingdom: St Mary’s Hospital, Imperial Healthcare NHS Trust, London (IHCT); Chelsea & Westminster Hospital NHS Foundation Trust, London (CWFT) and the John Radcliffe Hospital, Oxford University Hospitals NHS Foundation Trust, Oxford (OUH). All participants consented to two nasopharyngeal swabs being taken. During this period the incidence of COVID-19 in the UK peaked.^21^

Paired samples collected from the same site in the same patient or staff member were tested in parallel POC and laboratory platforms, with results from CovidNudge testing reported before laboratory results were available. Smaller caliber (pediatric) swabs were used to insert into the CovidNudge cartridge, most commonly a flexible minitip FLOQswab™ (COPAN Diagnostics Inc, Italy), whilst a second parallel combined oropharyngeal and nasopharyngeal swab was collected using a standard swabs and placed in viral transport medium for processing in a central laboratory as per local protocols (**SUPPLEMENTARY METHODS**).

Laboratory samples were processed at United Kingdom Accreditation Service (UKAS) laboratories. Samples collected at CWFT and IHCT were processed at the North West London Pathology Laboratory (NWLP, Charing Cross Hospital). Those collected OUH were processed at the John Radcliffe Hospital. Assessment took place at the peak of the epidemic in the UK and performance was compared to the platform in use at the time of collection in local laboratories (**SUPPLEMENTARY METHODS**). Centralized laboratory testing and POC testing were performed by separate staff members. Staff performing centralized laboratory testing were blinded to the POC test results and vice-versa.

Samples were collected from three groups: i) **Group 1** - self-referred, non-hospitalized healthcare workers or their family members with suspected COVID-19 (10th April to 12th may) at two sites (ICHT, OUH); ii) **Group 2** - patients admitted to emergency department with suspected COVID-19 at one site (ICHT). Suspected COVID-19 was defined as a patient presenting with any of the following: temperature ≥37.8°C; clinical evidence of pneumonia (e.g. cough, dyspnoea); hypoxia or an abnormal chest radiograph. Hospital staff were encouraged to self-refer and were eligible for testing if they self-reported any of the following symptoms: Fever ≥37.8°C or subjective fever, fatigue or malaise, cough and/or sputum production, muscle aches, headache, sore throat, profound loss of smell and taste. iii) **Group 3 -** consecutive hospital inpatient admissions with or without suspected COVID-19 from 12^th^ to the 18th May at one site (CWFT).

### Approvals

Participants in Group 2 were consented as part of the communicable disease research tissue bank (ethical approval ref 15/SC/0089). Following derogation from the Medicines and Healthcare Regulatory Agency (MHRA) evaluation within staff testing at all three sites was performed as a service evaluation in parallel with routine SARS-CoV-2 RT-PCR testing. Verbal or written consent for an additional swab was obtained from each participant and results from POC testing were not fed back to the individual participants. Analysis of Group 3 was conducted as a service evaluation approved by the Point of Care Committee at Chelsea & Westminster NHS Foundation Trust and results were used to inform patient care.

### Statistical Analyses

Data analysis was performed using R version 4.0^22^ using the epiR^23^ and the pheatmap^24^ packages. The primary analysis was conducted on paired samples collected on the same day. A secondary analysis was performed by sub-group, including by sample month, study site, location of sampling and comparator platform. Samples testing invalid on the CovidNudge platform were not included in the primary sensitivity analysis and were analysed separately. One batch of eight samples collected on one day at one site were excluded due to laboratory assay failure.

## 6. RESULTS

In vitro analysis with spiked SARS-CoV-2 RNA (**SUPPLEMENTARY METHODS**) found the lower limit of detection (LLOD) to be 5 viral RNA copies/ul for the *n3* assay, 10 viral RNA copies/ul for *n1, n2* and *E* assays whilst LLOD for *rdrp1*, *rdrp2* and *n1* targets was 50 viral RNA copies/ul (Supplementary Table 1Error! Reference source not found.). When the cartridge was spiked with whole virus particles into the lysis buffer chamber, the lower limit of detection was 1×10^4^ viral particles/sample for the n1, *n2* and *n3* targets (Supplementary Table 2).^5^

Clinical assessment was performed over a six-week period between the 2^nd^ April and 18^th^ May 2020. A total of 449 same-day samples were collected. Complete clinical data, paired with laboratory tests were available for 386, which were included in the primary analysis. The median age of study participants was 46 years (interquartile range 31 to 66 years) and 68% were female. A total of 280/386 (73%) of samples were collected from Group 1, 15/386 (4%) from Group 2 and 91/386 (23%) from Group 3 (**Figure 2**).

**Figure 2.**
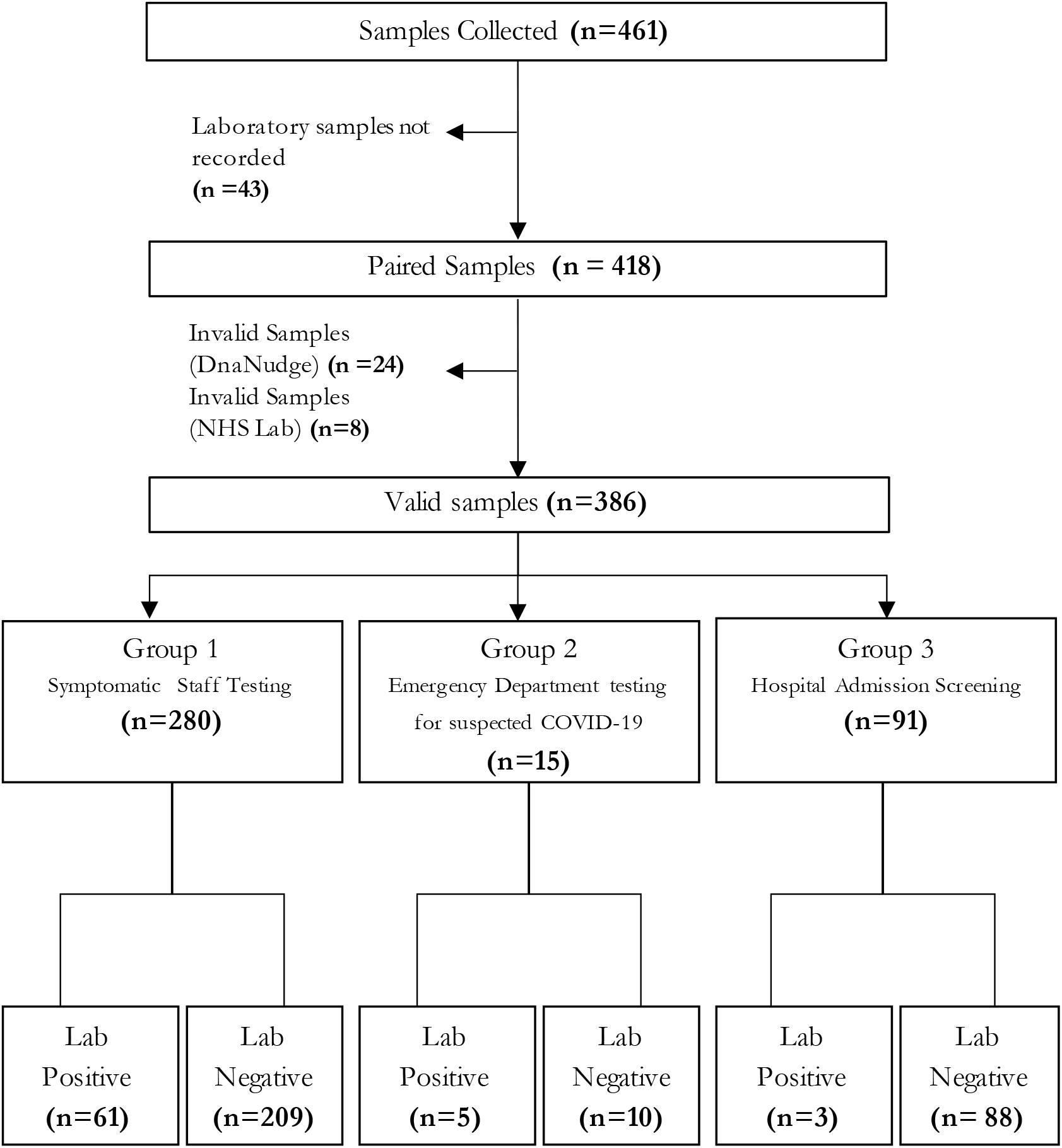
Profile of clinical study. Tests were considered valid if ≥3/6 replicates of RNaseP amplified. Suspected COVID-19 in the emergency department was defined as a patient presenting with any of the following: temperature =>37.8°C; clinical evidence of pneumonia (e.g. cough, dyspnoea); hypoxia or an abnormal chest radiograph. Healthcare workers were eligible for testing if they self-reported any of the following symptoms: Fever =>37.8 C or subjective fever, fatigue or malaise, cough and/ or sputum production, muscle aches, headache, sore throat, profound loss of smell and taste.

**Figure 3.**
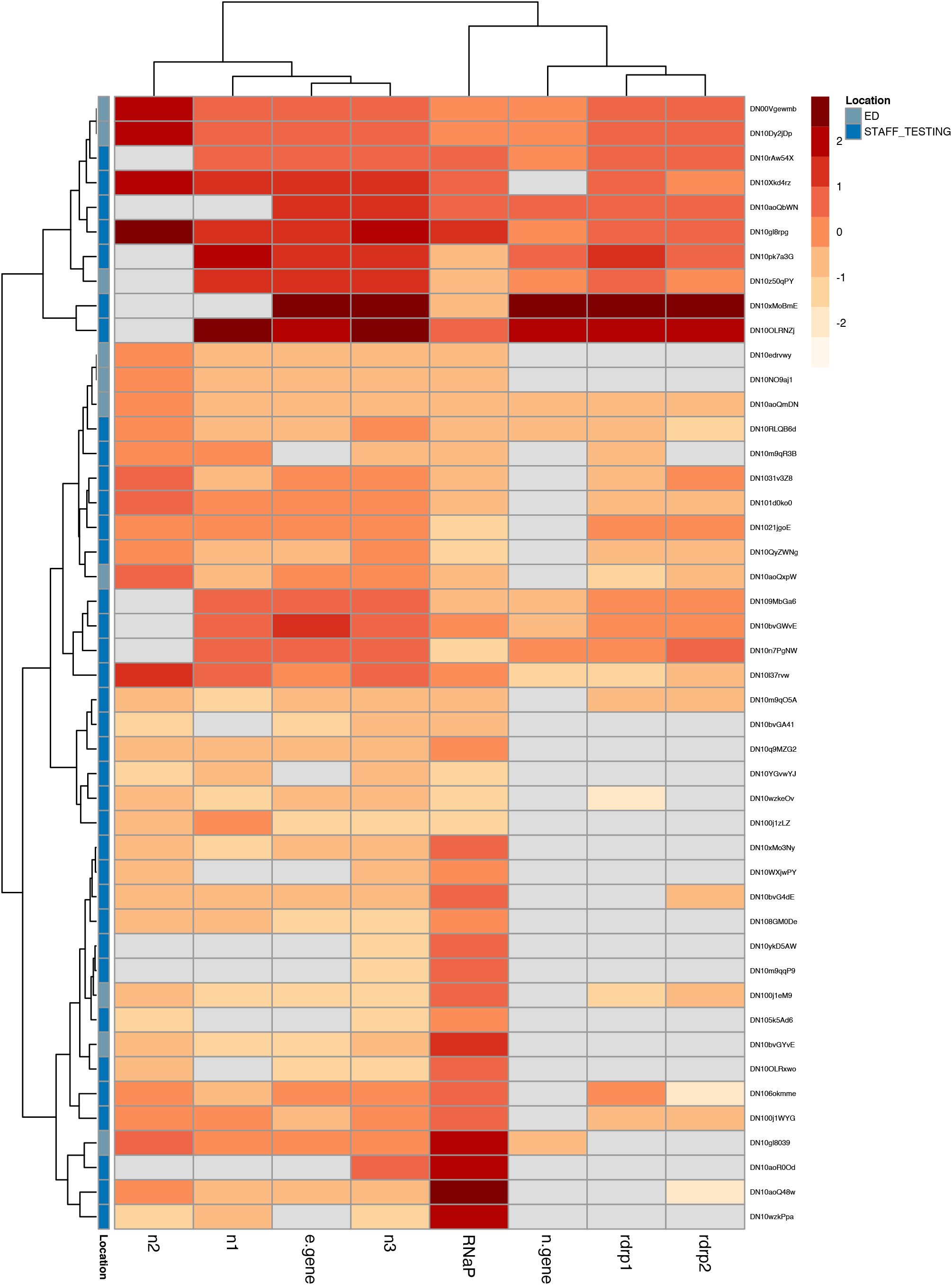
Heat map illustrating SARS-CoV-2 gene targets amplified in the CovidNudge point of care test. Rows correspond to samples and columns correspond to target genes spotted on the amplification unit of the CovidNudge cartridge. Illustrated are samples testing positive on the point of care platform, corresponding to samples where ≥2 replicates of at least one viral gene target amplified with ≥3 human RNaseP control replicates amplifying. Displayed are mean c values for each gene target amplified, corresponding to the inflection point of the sigmoid reaction curve, where the PCR reaction stops its exponential growth phase and begins its exponential decay phase.20 The c values are scaled using min-max normalisation, with a higher score corresponding to lower cycle number. Targets not amplified are displayed in grey. Clustering by Euclidean distance.

**Table 1.** Clinical assessment of point of care testing. Presented are paired samples collected contemporaneously. Samples testing invalid on the point of care test are not included (n=23). Results are presented according to location of testingg, context of testingg, laboratory platform and time period of testing. All samples were collected by nasopharyngeal swabs.

|  |  | Laboratory Testing |  | Point of Care Testing |  |  |  |  |  |  |  |
| --- | --- | --- | --- | --- | --- | --- | --- | --- | --- | --- | --- |
|  | Tested (n) | Positive | Negative | Positive | Negative | Prevalence | Sensitivity<br>(95% CI) | Specificity<br>(95% CI) | Positive<br>Predictive<br>Value<br>(95% CI) | Negative<br>Predictive<br>Value<br>(95% CI) | Negative<br>Likelihood<br>Ratio<br>(95% CI) |
| **Total** | 386 | 71 | 315 | 67 | 319 | 0.18 (0.15,<br>0.23) | 0.94 (0.86,<br>0.98) | 1.00 (0.99,<br>1.00) | 1.00 (0.94,<br>1.00) | 0.99 (0.97,<br>1.00) | 0.06 (0.02,<br>0.15) |
| **Sample context** |  |  |  |  |  |  |  |  |  |  |  |
| *Symptomatic Staff Testing* | 280 | 61 | 209 | 57 | 213 | 0.23 (0.18,<br>0.28) | 0.93 (0.84,<br>0.98) | 1.00 (0.98,<br>1.00) | 1.00 (0.94,<br>1.00) | 0.98 (0.95,<br>0.99) | 0.07 (0.03,<br>0.17) |
| *Emergency Department* | 15 | 5 | 10 | 5 | 10 | 0.33 (0.12,<br>0.62) | 1.00 (0.48,<br>1.00) | 1.00 (0.69,<br>1.00) | 1.00 (0.48,<br>1.00) | 1.00 (0.69,<br>1.00) | 0.00 (0.00,<br>NaN) |
| *All Hospital admissions* | 91 | 3 | 88 | 3 | 88 | 0.03 (0.01,<br>0.09) | 1.00 (0.29,<br>1.00) | 1.00 (0.96,<br>1.00) | 1.00 (0.29,<br>1.00) | 1.00 (0.96,<br>1.00)) | 0.00 (0.00,<br>NaN) |
| **Sample period** |  |  |  |  |  |  |  |  |  |  |  |
| *April 2020* | 272 | 68 | 204 | 64 | 208 | 0.25 (0.20,<br>0.31) | 0.94 (0.86,<br>0.98) | 1.00 (0.98,<br>1.00) | 1.00 (0.94,<br>1.00) | 0.98 (0.95,<br>0.99) | 0.06 (0.02,<br>0.16) |
| *May 2020* | 114 | 3 | 111 | 3 | 111 | 0.03 (0.01,<br>0.07) | 1.00 (0.29,<br>1.00) | 1.00 (0.97,<br>1.00) | 1.00 (0.29,<br>1.00) | 1.00 (0.97,<br>1.00) | 0.00 (0.00,<br>NaN) |

The overall prevalence of laboratory positive tests was 18% (71/386) with the highest prevalence being observed in patients attending the emergency department with suspected COVID-19 (33%; 95% CI 12-62%) and in samples collected in the month of April 2020 (25%; 95%CI 20-31%). The prevalence was lower in staff testing (group 1; 23% [18-28%]) and inpatient screening (group 3; 3% [1-9%]). In the primary analysis, the overall sensitivity of the POC test compared with a laboratory-based testing was 94% (95% Confidence interval 86-98%) with a specificity of 100% (99-100%; positive predictive value [PPV] 100% [94-100%]; negative predictive value [NPV] 99% [97-100%]) (**Table**). The platform performed equally well when compared against a range of laboratory-based platforms and in different clinical settings (**Supplementary Table 3**).

A subset of samples collected from symptomatic staff testing in one site (102/386; 26%) were run on three RT-PCR platforms (the CovidNudge point-of care test, the Public Health England RT-PCR assay targeting *rdrp* and the ThermoFisher assay targeting *orf1ab*, the spike [S] gene and the nucleocapsid [N] gene - **SUPPLEMENTARY METHODS**). Of these, 78/102 (76%) tested negative on all three platforms. Of samples testing positive with at least one assay (24/102 [24%]), a total of 22/24 (92%) were congruent across all three assays (**Supplementary Figure 2**).

The viral targets amplified varied markedly between individuals, with the most common amplified targets being the *n*3, *e-* and *n1* targets (Error! Reference source not found.Error! Reference source not found.).

Twenty-four samples processed on the point of care platform were reported as invalid due to failure to amplify human *RNaseP* in the point of care test, of which 22 had corresponding results from a laboratory specimen; of these, 16/22 (73%) tested negative.

## 7. DISCUSSION

In a diagnostic accuracy study conducted during the first peak of the UK COVID-19 pandemic, we have demonstrated that a lab-free point-of-care diagnostic test for SARS-CoV-2 had 94% sensitivity and 100% specificity when compared with standard laboratory-based RT-PCR. The key advantage of the CovidNudge platform is as a fully-automated direct sample-to-answer platform, removing the need for the laboratory infrastructure required for traditional RT-PCR. The run-time (under 90 minutes) is more rapid than other laboratory based diagnostic platforms.^9,13^ The data suggest that the CovidNudge platform has comparable or greater sensitivity and specificity than other rapid assays using dry swabs,^15,25^ and this will require head to head evaluation in future. In contrast to other rapid tests which still require viral transport medium and a simple sample transfer step,^15^ swabs are loaded directly into a fully sealed cartridge which allows safe testing outside of laboratory, potentially including primary care and community settings. We acknowledge that accuracy and a rapid run-time represents only some aspects of real-world POC test deployment. Prospective effectiveness studies are required to assess operational challenges, including access to equipment, impact on clinical decision making, cost effectiveness and equity of access.

The cartridge design allows the inclusion of multiple assays. One of these, human *RNaseP* control, is able to help ensure sample adequacy, a major challenge with many existing assays which cannot distinguish a true negative from an insufficient sample. In our study, 73% of samples reported as invalid on the POC platform (due to negative control) were reported as negative on laboratory assays lacking a sample adequacy control, some of which may have been false negatives. Reporting invalid results rapidly allows clinical decision makers the opportunity to repeat a test where the information is needed for clinical management. At the onset of the epidemic, the inclusion of several validated assays for different viral targets was expected to improve sensitivity. Surprisingly, one target in the N gene (*n3*) was positive in all positive cases, whereas *rdrp1* and *rdrp2* targets performed less well, consistent with previous reports.^26^ The design of the cartridge (**Figure 1C**), with each assay distributed across the analytical unit, means this difference is more likely due to biological differences in assay performance, than technical performance of the cartridge. Future adaptation will be to replace redundant assays with targets for respiratory syndromic screening (e.g. influenza, RSV) in anticipation of the diagnostic challenges on entering annual influenza season. Further work is required to understand how the algorithm relates to standard PCR measurements, such as the cycle threshold (C_t_) value, as well as virus viability, viral load, transmissability, and the performance of sgRNA targets in the cartridge to assess infectivity.^5,27^

We acknowledge the limitations of our study. The clinical assessment took place during a period of exceptionally heavy demand on clinical and laboratory services in the UK. It was not possible to use a single laboratory platform for comparison as the supply of reagents was inconsistent and unpredictable. Cross-platform comparison of two laboratory platforms was performed in a subset of samples. Given the POC assay had comparable performance against a range of other commercial platforms run in different labs, it is reasonable to expect that similar performance would be observed in different clinical settings. Following recent CE marking to allow testing outside of hospitals, and NHS procurement, a standard process for the roll-out is being developed by the NHS taking into account this issue. Nevertheless, we advocate for local assessment to compare performance against existing local standards of care when the device is first deployed in a new setting. Falling incidence of infection during the period of study meant it was not possible to validate the test with a larger number of positive samples, however, the high specificity in a cohort with low background prevalence is reassuring given the risks of incorrectly placing a patient without infection into a ward designated for SARS-CoV-2 infected patients.

Centralised testing with RT-PCR has the advantage of high throughput processing that cannot be achieved by the CovidNudge platform at the current time. As each processing unit (NudgeBox) can process one cartridge at a time, the assay has relatively low throughput and multiple processing units maybe required depending on the clinical setting. However, judicious application of point-of-care tests could relieve the burden on central laboratories and increase overall testing capacity, complementing existing approaches. The platform has a role in testing strategies where results can impact real-time decision making such as prescribing specific SARS-CoV-2 therapy (e.g. remdesivir or dexamethasone), triaging unscheduled admissions (e.g. to emergency departments and maternity units) and screening elective admissions or staff (e.g. prior to procedures such as surgery or chemotherapy). In addition, each device is linked to a secure cloud-based database via WiFi, allowing results to be delivered directly to clinical information systems. The potential exists to link to patient smartphones applications and/or test and trace facilities, although further work on acceptability, privacy and information governance are planned for future work. In principle, the platform is well suited to testing in primary care and community settings (e.g. long term care facilities or contact tracing programs) with potential for use in non-healthcare settings (e.g. prisons, transport hubs or offices). However, further studies of real-world effectiveness in non-clinical settings would be required prior to widespread deployment.

Enhanced testing forms a central pillar of global efforts to control SARS-CoV-2.^27^ We have described the first report of the development and clinical assessment of highly sensitive and specific rapid point-of-care platform for the detection of SARS-CoV-2, validated in frontline clinical settings during the first peak of the COVID-19 pandemic. The device is already in use in clinical settings in the UK and is one component of the testing strategy which is required to contain the COVID19 pandemic.^28^

## Data Availability

De-identified study data are available from the corresponding author upon reasonable request.

## ACKNOWLEDGEMENTS

The authors wish to acknowledge the contribution of participants and the staff of Imperial College NHS Trust, Oxford University NHS Trust, North West London Pathology and Imperial College Molecular Diagnostic Unit. The work was supported by the Biomedical Research Centre of Imperial College NHS Trust. M.M.G. is supported in part by the NIHR Imperial Biomedical Research Centre. GC is an NIHR Research Professor and Investigator within the NIHR London In-vitro Diagnostic Collaborative. Part of this work was supported by the National Institute for Health Research Health Protection Research Unit (NIHR HPRU) in Healthcare Associated Infections and Antimicrobial Resistance at Oxford University in partnership with Public Health England (PHE) [grant HPRU-2012-10041] and the NIHR Biomedical Research Centre, Oxford. The views expressed in this publication are those of the authors and not necessarily those of the NHS, the National Institute for Health Research, the Department of Health or Public Health England.

## AUTHOR CONTRIBUTIONS

Assay design and development was performed by CT (genetics and bioengineering design), RS (assay development and molecular biology), MS (platform technology and cartridge design), MK (genetics and microbiology), TH, SDM, FL, JB and AO.

Laboratory development was supported by WB, GPT and GC.

Clinical evaluation was led by MMG, JB-N, BF, RK, GD, LSPM, NM, KJ,DC, GC and AB.

NHS laboratory testing was undertaken by IKS, PR, DM DC and KJ.

Analysis was performed by MMG, CT, GC, RS and MS.

The first draft of manuscript was written by MMG and GC. All authors reviewed and approved the final manuscript.

CT is the co-inventor of the DnaNudge CovidNudge system.

## COMPETING INTEREST DECLERATION

CT, RS, MS, CI, MK, TH, SDM, FL, JB and AO are employees of DnaNudge. CT is named on the patent for method and apparatus for analyzing biological specimens on the DnaNudge platform (US Patent No: US 10,093,965 B2^16^. LSPM has consulted for bioMerieux (2013-2020), DNAelectronics (2015), Dairy Crest (2017–2018), Pfizer (2018-2020), and Umovis Lab (2020), received speaker fees from Profile Pharma (2018), received research grants from the National Institute for Health Research (2013–2019), Leo Pharma (2016), and CW+ Charity (2018-2019), and received educational support from Eumedica (2016–2017). NM has received speaker fees from Beyer (2016) and Pfizer (2019) and received educational support from Eumedica (2016) and Baxter (2017). All other authors have no conflicts of interest to declare.

## 9. FIGURE LEGENDS

**Supplementary Figure 1.**
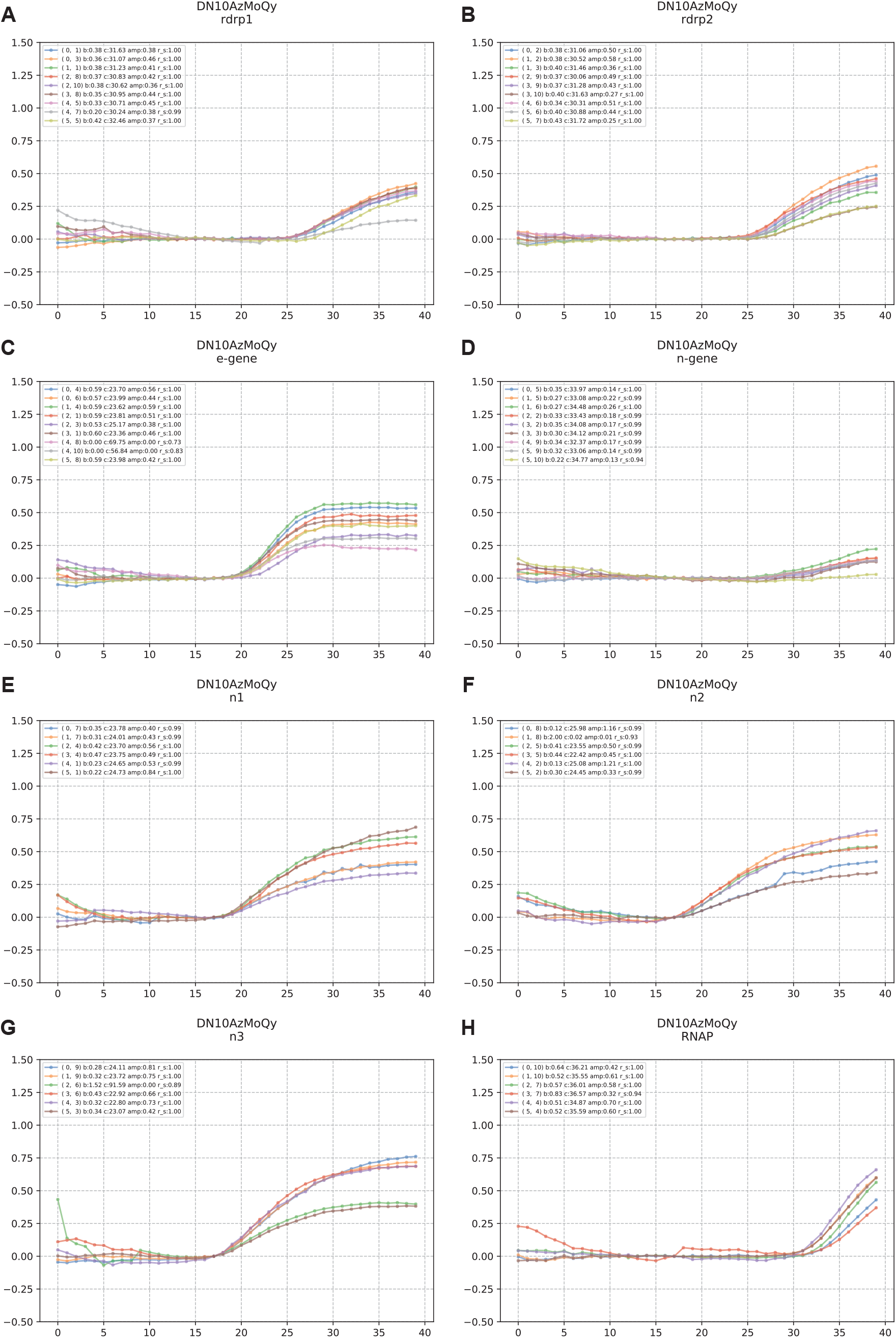
Representative reaction curve plots for strongly positive SARS-CoV-2 PCR Displayed are reaction curves for target genes (A) rdrp1; (B) rdrp2; (C) e-gene; (D) n-gene; (E)n1; (F) n2 and (G) n3. in the assay with accompanying technical replicates. X-axis corresponds to cycle number and Y-axis corresponds to fluorescence index. For a well to be classified as having amplified, the data should reflect the exponential growth and decay of a PCR reaction and be “sigmoid-like” according to a pre-defined algorithm (see Supplementary Methods).

**Supplementary Figure 2.**
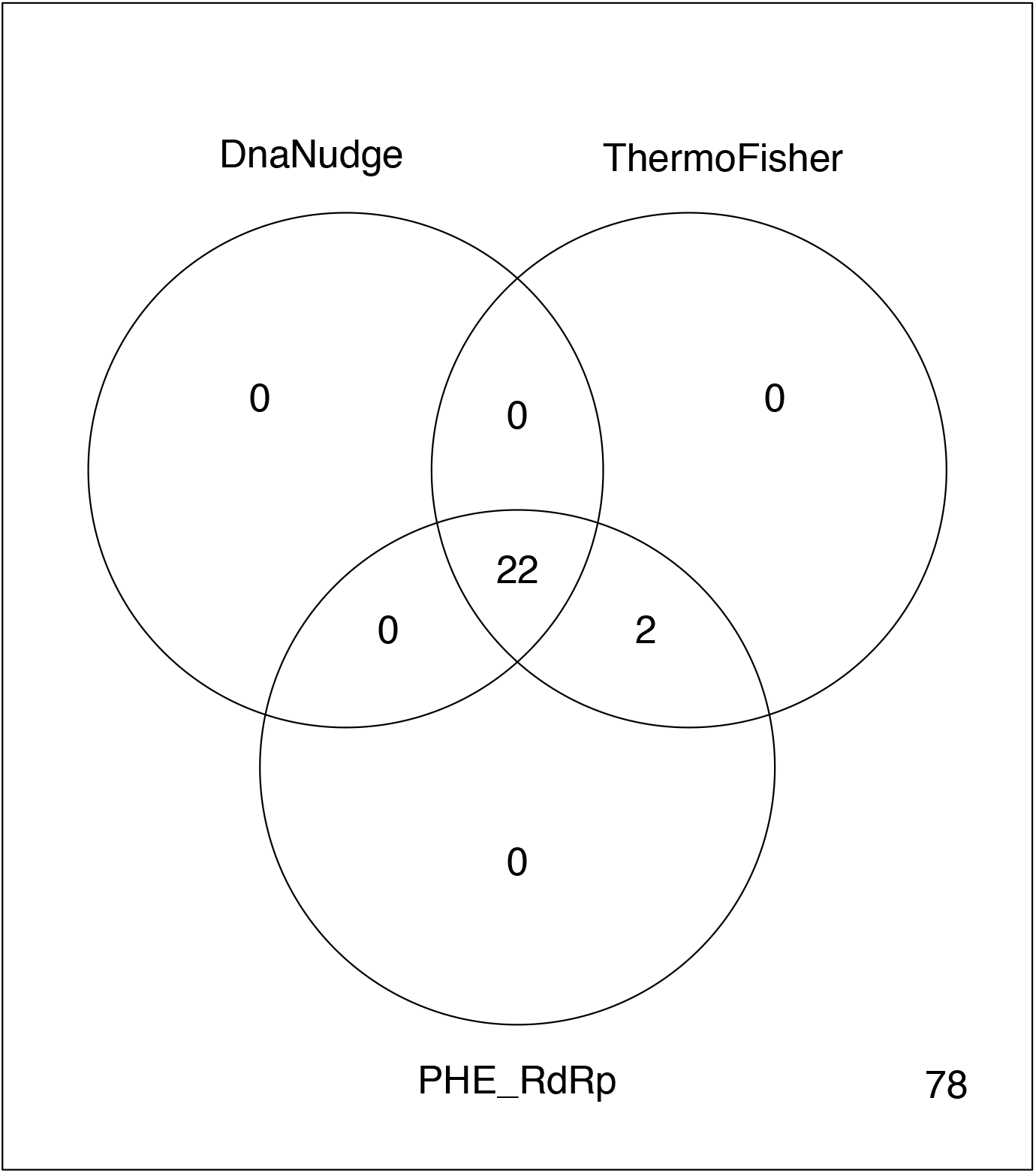
Venn diagram illustrating cross assay assessment. One hundred and two samples (all collected at the John Radcliffe Hospital, Oxford) were run concurrently on three RT-PCR assays. Numbers displayed in overlapping circles represent the number of positive tests per labelled assay

## 13. SUPPLEMENTARY METHODS

### Laboratory Testing

The Public Health England assay is a real-time RT-PCR assay targets a 100bp fragment from a conserved region of the SARS-CoV-2 RNA-dependent-RNA polymerase (RdRp).29 The AusDiagnostics (New South Wales, Australia) assay is a multiplex-tandem polymerase chain reaction (MT-PCR) targeting the conserved region of *Orf1ab* and *Orf8* from the SARS-CoV-2 genome. RNA extraction was undertaken using the Qiagen EZ1 or the AusDiagnostics MT-Prep kit. Samples process at the Imperial Molecular Diagnostics Unit SARS-CoV-2 were analyzed using real-time quantitative PCR monitored by a FAM-conjugated probe in a BioRad (California, United Statea) CFX Real Time PCR system, using *E*-gene primers and probe reported as reported by Corman et al.7 RNA extracted from patient samples was carried out on the Felix liquid handling robot and amplified using real-time quantitative PCR monitored by a FAM-conjugated probe in a BioRad CFX Real Time PCR system. The Roche (Basel, Switzerland) assay is a is a dual-target real-time RT-PCR assay targeting conserved regions *Orf1ab* and *E*-genes, processed on the Roche 6800 platform. The Abbott (Illinois, United States) real-time SARS-CoV-2 assay is a dual-target real-time RT-PCR assay targeting conserved regions of the RdRp and N-genes, run on an Abbott M2000 machine.19 The ThermoFisher (Massachusetts, United States) assay is a multiplex real-time RT-PCR with primers and probes targeting *orf1ab*, the spike (*S*) gene and the nucleocapsid (*N*) gene, as well as incorporating an *RNaseP* control. Extraction was undertaken on a FeliX liquid handling robot with amplification on an Analytik Jena qTower.

### CovidNudge protocol

To start a test, the user scans the cartridge barcode using the capsule. After placing the cartridge into the NudgeBox and the Capsule on its top lid on the NudgeBox, the user can start the test by pressing the Capsule button. The Capsule informs the NudgeBox of the DnaCartridge barcode, and the NudgeBox communicates that with the DnaNudge cloud to register a new test or abort in case it is an invalid barcode (e.g. previously used).

The test starts with moving the lysis buffer to the swab chamber by rotating and depressurising/pressurising the mixing chamber between lysis and swab chambers. The lysis kills and deactivates the viral sample and releases the sample RNA. Silica frit filters are mounted on to the port in the mixing chamber which can capture RNA molecules. By moving back the lysis buffer from the swab chamber to the mixing chamber, the extracted RNA strands bind to the silica frit. In the next step, wash buffer is passed through the mixing chamber and the frit to remove any debris. In the third step, the elution buffer releases the RNA strands from the frit. By turning the mixing chamber toward the master mix chamber, the elution buffer containing the sample RNA is used to reconstitute the lyophilised RT master mix. This action is repeated to create a homogenous mix. In the last step of sample preparation, mixing chamber turns toward the AU filling port of the SPU to fill the AU.

Once the AU is filled, the NudgeBox clamps the AU between lower and upper thermal subsystems. This helps with filling the reaction wells and ensures that there is no carry over between any two neighbouring wells. The RT-PCR starts with a reverse transcriptase step at 45°C for 5 minutes, a 2-minute RT inactivation and Taq activation step at 95°C, followed by 40 cycles of PCR (3-second denaturation steps at 95°C and 30-second annealing/extension steps at 60°C). At the end of every annealing/extension step, the imaging system measures the light intensity of every reaction well.

Exposed surfaces of the instrument are cleaned regularly between operators with 10% bleach, followed by an IPA wipe to remove any residual bleach. Following the test, the cartridge is disposed of following standard laboratory disposal procedures.

### Primers and Probes

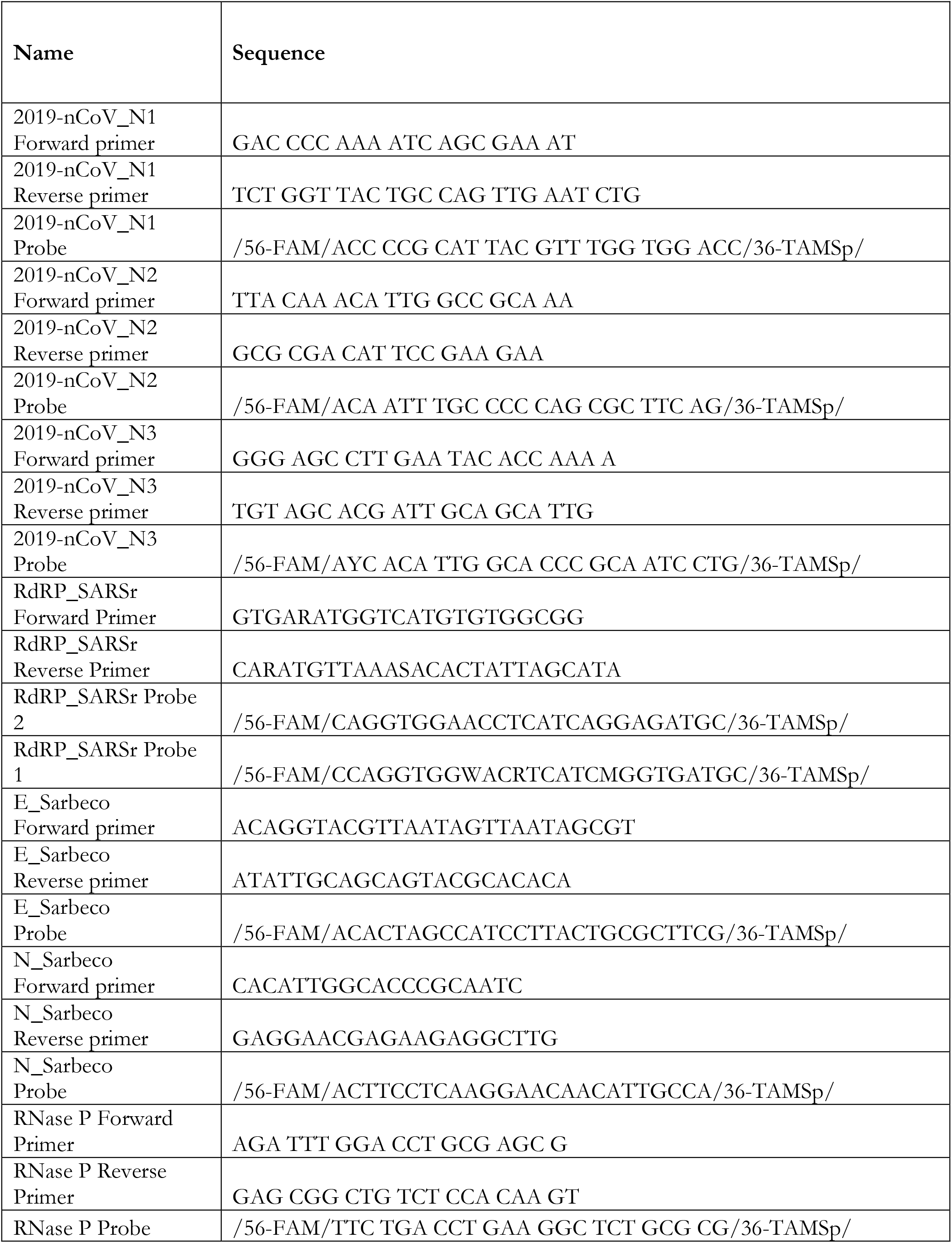

Exclusivity of the assays with respect to the Coronaviridae family was evaluated in silico by mapping the primer and probe sequences to homologous sequences downloaded from the NCBI database. The WHO N-gene, WHO E-gene and WHO RdRp-1 assays are predicted to detect human SARS-coronavirus and bat SARS-like coronaviruses in the subgenus Sarbecovirus. CDC N3 assay may also detect Sarbecovirus other than SARS-CoV-2. No cross-reactivity with human coronaviruses OC43, HKU1, NL63, 229E or MERS-coronavirus was detected for any assays. NCBI primer-BLAST tool was used to assess potential cross-reactivity with other respiratory pathogens and high-priority organisms. No unintended cross-reactivity was detected for any organisms listed below:

| Organism |  |
| --- | --- |
| Adenovirus A/B/C/D/E | *Haemophilus influenzae* |
| Enterovirus A/B/C | *Legionella* |
| Human metapneumovirus | *Leptospira* |
| Influenza A/B/C | *Moraxella catarrhalis* |
| Parainfluenza virus 1-4 | *Mycobacterium tuberculosis* |
| Parechovirus | *Mycoplasma pneumoniae* |
| Respiratory syncytial virus | *Neisseria elongate* |
| Rhinovirus A/B | *Neisseria meningitidis* |
| *Bacillus anthracis* | *Pneumocystis jirovecii* |
| *Bordetella pertussis* | *Pseudomonas aeruginosa* |
| *Candida albicans* | *Staphylococcus aureus* |
| *Chlamydia pneumoniae* | *Staphylococcus epidermidis* |
| *Chlamydia psittaci* | *Staphylococcus salivarius* |
| *Corynebacterium diphtheriae* | *Streptococcus pneumoniae* |
| *Coxiella burnetii* | *Streptococcus pyogenes* |

### PCR analysis

Analysis from individual wells is subdivided into model fitting, post-processing and classification stages. The data is modelled by the following formula:

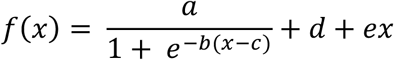

Where x is the PCR cycle. The first term, consisting of parameters *a, b*, and *c* (“the sigmoid term”) describe the exponential growth and decay in fluorescence intensity during a test. Parameters *d* and *e* account for system nonidealities, inter-test and inter-instrument variability. Raw data is fitted to the model with least- squares curve fitting techniques which provides estimates for parameters *a, b* and *c*. To ensure well to well and test to test consistency, data from each well undergoes drift correction and normalisation. Using the model parameters calculated previously the data is re-simulated with the ***e*** parameter set to zero and multiplying the remaining terms by a normalisation factor.

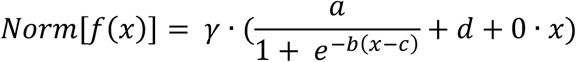

Where *γ* is the normalisation factor.

For a well to be classified as having amplified, the data should reflect the exponential growth and decay of a PCR reaction, simply, it should be “sigmoid-like”. This implies that the model parameters should fall within appropriate ranges. Specifically, inspection of the ***b*** and ***c*** parameters and the synthesis of two additional parameters (normalised sigmoid amplitude and r^2^) allow the algorithm to classify data as “sigmoid-like” or otherwise.

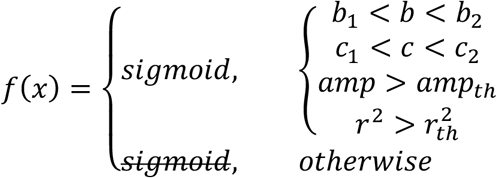

Where *b*_1_, *b*_2_, *c*_1_*, c*_2_ are upper and lower bounds for ***b*** and ***c*** respectively while *amp_th_* and 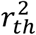 are thresholds over which the normalised amplitude and goodness of fit must exceed.

To identify the DC line and avoid the initial cycles noise, a median value of a range of midway cycles is used to normalise and adjust the base line. This adjustment is to help with applying standardised rules on calling a signal a positive or negative. When the sigmoid fit is applied and passes an r-squared criteria to indicate if the signal could be properly modelled with sigmoid or not, the parameters helping with that fit are compared against normal signal threshold values.

Initial analysis of samples collected in the emergency department at St Mary’s hospital between the 2nd to 19th April were performed by manual inspection of amplification curves by laboratory staff blinded to the results of centralized laboratory testing, assessing for the inflection point of the sigmoid reaction curve. Subsequent analysis was performed algorithmically. By running the optimized algorithm on the manual data before the 19th April on samples run against the AusDiagnostic platform, estimates sensitivity is reduced. However, this would be expected since limits of detection on a nested PCR are a lot higher than a standard RT-PCR.

### Limits of detection

In order to measure the limits of detection (LOD) of assay of the CovidNudge platform, control RNA (Twist Synthetic SARS-CoV-2 RNA Controls, Twist Biosciences, USA) was diluted to varying concentrations and spiked in the RT-PCR lyophilised master mix chamber of the cartridges. Promega human DNA(~0.6ng/ul) was also spiked and used as a carrier RNA to prevent any low label RNA binding to plastic. Analysis was also performed using viral particles a known concentration (1×10^6 copies per mL) in a proprietary matrix were purchased from ZeptoMetrix, Product Desription: NATtrol SARS-Related Coronavirus 2(SARS-CoV-2), Stock, Cat No: NATSARS(COV2)-ST). Samples were processed as outlined in the CovidNudge protocol and PCR analysis sections above.

**Supplementary Table 1.** Assessment of lower limit of detection for CovidNudge platform. Control RNA (Twist Synthetic SARS-CoV-2 RNA Controls, Twist Biosciences, USA) was diluted to varying concentrations and spiked in the RT-PCR lyophilised master mix chamber of the cartridges. Displayed are mean c values for each gene target amplified, corresponding to the inflection point of the sigmoid reaction curve, where the PCR reaction stops its exponential growth phase and begins its exponential decay phase.

| Total viral RNA input (copies) | Conc. In reaction (RNA copies/ul) |  | Target Gene |  |  |  |  |  |  |
| --- | --- | --- | --- | --- | --- | --- | --- | --- | --- |
|  |  |  | RdRP1<br>(Replicates = 9) | RdRP2<br>(Replicates = 9) | E_Sarbeco<br>(Replicates = 9) | N_Sarbeco<br>(Replicates = 9) | N1<br>(Replicates = 6) | N2<br>(Replicates = 6) | N3<br>(Replicates = 6) |
| 50,000 SARS-CoV-2 | 125 | *Number of Replicates Amplified* | 8/9 | 6/9 | 9/9 | 2/9 | 6/6 | 6/6 | 6/6 |
|  |  | *Mean Cycle Threshold* | 34 | 34 | 30 | 36 | 31 | 31 | 31 |
|  |  | *End Point Fluorescence* | 0.35 | 0.35 | 0.8 | 0.2 | 0.7 | 0.6 | 0.7 |
| 50,000 SARS-CoV-2 | 125 | *Number of Replicates Amplified* | 5/9 | 6/9 | 9/9 | 3/9 | 6/6 | 6/6 | 6/6 |
|  |  | *Mean Cycle Threshold* | 35 | 33 | 31 | 36 | 32 | 30 | 30 |
|  |  | *End Point Fluorescence* | 0.4 | 0.65 | 0.9 | 0.25 | 0.85 | 1.1 | 1 |
| 50,000 SARS-CoV-2 | 125 | *Number of Replicates Amplified* | 3/9 | 4/9 | 7/9 | 2/9 | 6/6 = | 6/6 | 6/6 |
|  |  | *Mean Cycle Threshold* | 36 | 35 | 33 | 37 | 31 | 32 | 32 |
|  |  | *End Point Fluorescence* | 0.25 | 0.35 | 0.3 | 0.15 | 0.65 | 0.4 | 0.55 |
| 40,000 SARS-CoV-2 | 100 | *Number of Replicates Amplified* | 4/9 | 6/9 | 9/9 | 3/9 = | 6/6 | 6/6 | 6/6 |
|  |  | *Mean Cycle Threshold* | 35 | 24 | 32 | 37 | 31 | 32 | 31 |
|  |  | *End Point Fluorescence* | 0.7 | 1 | 1 | 0.35 | 1 | 0.7 | 0.9 |
| 40,000 SARS-CoV-2 | 100 | *Number of Replicates Amplified* | 3/9 | 5/9 | 9/9 | 2/9 | 5/6 | 6/6 | 6/6 |
|  |  | *Mean Cycle Threshold* | 37 | 35 | 31 | 36 | 31 | 31 | 31 |
|  |  | *End Point Fluorescence* | 0.45 | 10.6 | 0.75 | 0.35 | 0.95 | 2 | 1.1 |
| 25,000 SARS-CoV-2 | 62.5 | *Number of Replicates Amplified* | 1/6 | 1/6 | 3/6 | 0/0 | 2/6 | 2/6 | 5/6 |
|  |  | *Mean Cycle Threshold* | 41 | 38 | 35 | - | 35 | 35 | 35 |
|  |  | *End Point Fluorescence* | - | - | - | - | - | - | - |
| 20,000 SARS-CoV-2 | 50 | *Number of Replicates Amplified* | 1/9 | 1/9 | 4/9 | 2/9 | 1/6 | 3/6 | 3/6 |
|  |  | *Mean Cycle Threshold* | 38 | 40 | 34 | 38 | 33 | 32 | 34 |
|  |  | *End Point Fluorescence* | - | - | - | - | - | - | - |
| 10,000 SARS-CoV-2 | 25 | *Number of Replicates Amplified* | No amplification | No amplification | 1/9 Ct 35 | No amplification | No amplification | 3/6 | 2/6 |
|  |  | *Mean Cycle Threshold* | - | - | 35 | - | - | 32 | 34 |
|  |  | *End Point Fluorescence* | - | - | - | - | - | - | - |
| 1000 SARS-CoV-2 | 10 | *Number of Replicates Amplified* | No amplification | No amplification | 3/9 | No amplification | 1/6 | 4/6 | 6/6 |
|  |  | *Mean Cycle Threshold* | - | - | 35 | - | 33 | 34 | 35 |
|  |  | *End Point Fluorescence* | - | - | - | - | - | - | - |
| 1000 SARS-CoV-2 | 5 | *Number of Replicates Amplified* | No amplification | 1/9 | No amplification | No amplification | No amplification | No amplification | 4/6 |
|  |  | *Mean Cycle Threshold* | - | 35 | - | - | - | - | 34 |
|  |  | *End Point Fluorescence* | - | - | - | - | - | - | - |
| 1000 SARS-CoV-2 | 2.5 | *Number of Replicates Amplified* | No amplification | No amplification | No amplification | No amplification | No amplification | No amplification | No amplification |
|  |  | *Mean Cycle Threshold* | - | - | - | - | - | - | - |
|  |  | *End Point Fluorescence* | - | - | - | - | - | - | - |

**Supplementary Table 2.**
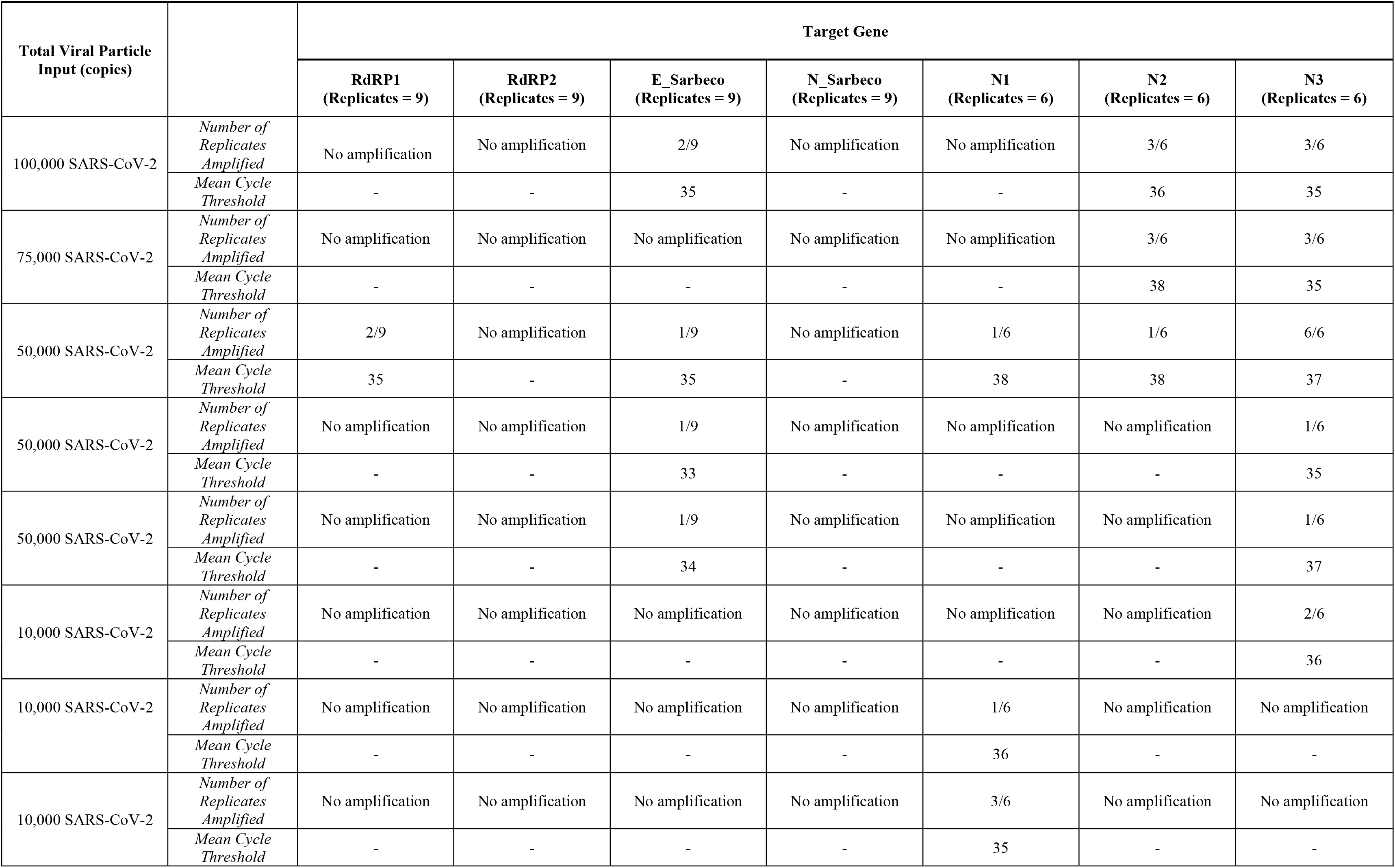
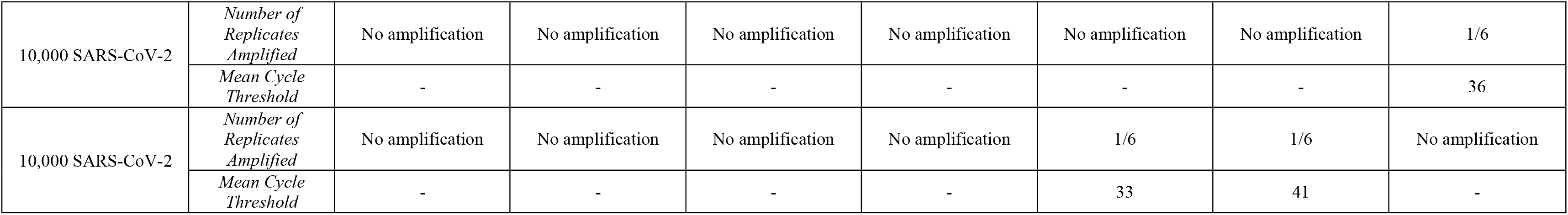
Assessment of lower limit of detection for CovidNudge platform using test SARS-CoV-2 viral particles. Control viral particles (Zeptometrix NATrol SARS-CoV-2 RNA Controls) was diluted to varying concentrations and spiked in the RT-PCR lysis chamber of the cartridges. Displayed are mean c values for each gene target amplified, corresponding to the inflection point of the sigmoid reaction curve, where the PCR reaction stops its exponential growth phase and begins its exponential decay phase.

| Total Viral Particle Input (copies) |  | Target Gene |  |  |  |  |  |  |
| --- | --- | --- | --- | --- | --- | --- | --- | --- |
|  |  | RdRP1<br>(Replicates = 9) | RdRP2<br>(Replicates = 9) | E_Sarbeco<br>(Replicates = 9) | N_Sarbeco<br>(Replicates = 9) | N1<br>(Replicates = 6) | N2<br>(Replicates = 6) | N3<br>(Replicates = 6) |
| 100,000 SARS-CoV-2 | Number of Replicates Amplified | No amplification | No amplification | 2/9 | No amplification | No amplification | 3/6 | 3/6 |
|  | Mean Cycle Threshold | - | - | 35 | - | - | 36 | 35 |
| 75,000 SARS-CoV-2 | Number of Replicates Amplified | No amplification | No amplification | No amplification | No amplification | No amplification | 3/6 | 3/6 |
|  | Mean Cycle Threshold | - | - | - | - | - | 38 | 35 |
| 50,000 SARS-CoV-2 | Number of Replicates Amplified | 2/9 | No amplification | 1/9 | No amplification | 1/6 | 1/6 | 6/6 |
|  | Mean Cycle Threshold | 35 | - | 35 | - | 38 | 38 | 37 |
| 50,000 SARS-CoV-2 | Number of Replicates Amplified | No amplification | No amplification | 1/9 | No amplification | No amplification | No amplification | 1/6 |
|  | Mean Cycle Threshold | - | - | 33 | - | - | - | 35 |
| 50,000 SARS-CoV-2 | Number of Replicates Amplified | No amplification | No amplification | 1/9 | No amplification | No amplification | No amplification | 1/6 |
|  | Mean Cycle Threshold | - | - | 34 | - | - | - | 37 |
| 10,000 SARS-CoV-2 | Number of Replicates Amplified | No amplification | No amplification | No amplification | No amplification | No amplification | No amplification | 2/6 |
|  | Mean Cycle Threshold | - | - | - | - | - | - | 36 |
| 10,000 SARS-CoV-2 | Number of Replicates Amplified | No amplification | No amplification | No amplification | No amplification | 1/6 | No amplification | No amplification |
|  | Mean Cycle Threshold | - | - | - | - | 36 | - | - |
| 10,000 SARS-CoV-2 | Number of Replicates Amplified | No amplification | No amplification | No amplification | No amplification | 3/6 | No amplification | No amplification |
|  | Mean Cycle Threshold | - | - | - | - | 35 | - | - |
| 10,000 SARS-CoV-2 | *Number of Replicates Amplified* | No amplification | No amplification | No amplification | No amplification | No amplification | No amplification | 1/6 |
|  | *Mean Cycle Threshold* | - | - | - | - | - | - | 36 |
| 10,000 SARS-CoV-2 | *Number of Replicates Amplified* | No amplification | No amplification | No amplification | No amplification | 1/6 | 1/6 | No amplification |
|  | *Mean Cycle Threshold* | - | - | - | - | 33 | 41 | - |

**Supplementary Table 3.** Assessment of point of care testing in paired samples collected contemporaneously presented by study site and comparator laboratory assay.

|  |  | Laboratory Testing |  | Point of Care Testing |  |  |  |  |  |  |  |
| --- | --- | --- | --- | --- | --- | --- | --- | --- | --- | --- | --- |
|  | Tested (n) | Positive | Negative | Positive | Negative | Prevalence | Sensitivity<br>(95% CI) | Specificity<br>(95% CI) | Positive<br>Predictive<br>Value<br>(95% CI) | Negative<br>Predictive<br>Value<br>(95% CI) | Negative<br>Likelihood<br>Ratio<br>(95% CI) |
| Site |  |  |  |  |  |  |  |  |  |  |  |
| *St Mary's Hospital, London* | 162 | 34 | 128 | 33 | 129 | 0.21 (0.15,<br>0.28) | 0.97 (0.85,<br>1.00) | 1.00 (0.97,<br>1.00) | 1.00 (0.89,<br>1.00) | 0.99 (0.96,<br>1.00) | 0.03 (0.00,<br>0.22) |
| *Chelsea & Westminster Hospital, London* | 91 | 3 | 88 | 3 | 88 | 0.03 (0.01,<br>0.09) | 1.00 (0.29,<br>1.00) | 1.00 (0.96,<br>1.00) | 1.00 (0.29,<br>1.00) | 1.00 (0.96,<br>1.00) | 0.00 (0.00,<br>NaN) |
| *John Radcliffe Hospital, Oxford* | 133 | 34 | 99 | 31 | 102 | 0.26 (0.18,<br>0.34) | 0.91 (0.76,<br>0.98) | 1.00 (0.96,<br>1.00) | 1.00 (0.89,<br>1.00) | 0.97 (0.92,<br>0.99) | 0.09 (0.03,<br>0.26) |
| Laboratory Assay |  |  |  |  |  |  |  |  |  |  |  |
| *AusDiagnostics_High-Plex* | 74 | 25 | 49 | 24 | 50 | 0.34 (0.23,<br>0.46) | 0.96 (0.80,<br>1.00) | 1.00 (0.93,<br>1.00) | 1.00 (0.86,<br>1.00) | 0.98 (0.89,<br>1.00) | 0.04 (0.01,<br>0.27) |
| *ROCHE* | 81 | 5 | 76 | 5 | 76 | 0.06 (0.02,<br>0.14) | 1.00 (0.48,<br>1.00) | 1.00 (0.95,<br>1.00) | 1.00 (0.48,<br>1.00) | 1.00 (0.95,<br>1.00) | 0.00 (0.00,<br>NaN) |
| *ABBOTT* | 66 | 4 | 62 | 2 | 64 | 0.06 (0.02,<br>0.15) | 0.50 (0.07,<br>0.93) | 1.00 (0.94,<br>1.00) | 1.00 (0.16,<br>1.00) | 0.97 (0.89,<br>1.00) | 0.50 (0.19,<br>1.33) |
| *ThermoFisher* | 21 | 0 | 21 | 0 | 21 | 0.00 (0.00,<br>0.16) | - | 1.00 (0.84,<br>1.00) | - | 1.00 (0.84,<br>1.00) | - |
| *Public Health England - RdRp* | 120 | 32 | 88 | 31 | 89 | 0.27 (0.19,<br>0.36) | 0.97 (0.84,<br>1.00) | 1.00 (0.96,<br>1.00) | 1.00 (0.89,<br>1.00) | 0.99 (0.94,<br>1.00) | 0.03 (0.00,<br>0.22) |
| *Imperial Molecular Diagnostics Unit* | 24 | 5 | 19 | 5 | 19 | 0.21 (0.05,<br>0.51) | 1.00 (0.48,<br>1.00) | 1.00 (0.82,<br>1.00) | 1.00 (0.48,<br>1.00) | 1.00 (0.82,<br>1.00) | 0.00 (0.00,<br>NaN) |

**Supplementary Table 4.** Secondary analysis. *Sensitivity and specificity when compared against all paired samples collected, including those not collected on the same date (n=47). Early validation samples for assessment of the point of care test in included samples collected from patients admitted to hospital with COVID-19 confirmed on nasopharyngeal swabs tested in a central laboratory. The median interval between sample collection for laboratory processing and point of care testing was 4 days (interquartile range 1 to7). ^†^All same-day samples including early validation samples collected prior to algorithm optimisation (n=10 valid samples ran on Abbott platform at Oxford).

|  |  | Laboratory Testing |  | Point of Care Testing |  |  |  |  |  |  |  |
| --- | --- | --- | --- | --- | --- | --- | --- | --- | --- | --- | --- |
|  | Tested (n) | Positive | Negative | Positive | Negative | Prevalence | Sensitivity (95% CI) | Specificity (95% CI) | Positive Predictive Value (95% CI) | Negative Predictive Value (95% CI) | Negative Likelihood Ratio (95% CI) |
| All samples collected including early validation samples* | 444 | 115 | 329 | 95 | 349 | 0.26 (0.22, 0.30) | 0.81 (0.72, 0.88) | 0.99 (0.98, 1.00) | 0.98 (0.93, 1.00) | 0.94 (0.91, 0.96) | 0.19 (0.13, 0.28) |
| Same day samples including early validation samples† | 394 | 75 | 319 | 70 | 324 | 0.18 (0.14, 0.22) | 0.91 (0.82, 0.96) | 0.99 (0.98, 1.00) | 0.97 (0.90, 1.00) | 0.98 (0.96, 0.99) | 0.09 (0.05, 0.19) |
| Sample Context |  |  |  |  |  |  |  |  |  |  |  |
| Symptomatic Staff Testing | 278 | 65 | 213 | 60 | 218 | 0.23 (0.19, 0.29) | 0.89 (0.79, 0.96) | 0.99 (0.97, 1.00) | 0.97 (0.88, 1.00) | 0.97 (0.93, 0.99) | 0.11 (0.05, 0.22) |
| Emergency Department | 15 | 5 | 10 | 5 | 10 | 0.33 (0.12, 0.62) | 1.00 (0.48, 1.00) | 1.00 (0.69, 1.00) | 1.00 (0.48, 1.00) | 1.00 (0.69, 1.00) | 0.00 (0.00, NaN) |
| All Hospital admissions | 91 | 3 | 88 | 3 | 88 | 0.03 (0.01, 0.09) | 1.00 (0.29, 1.00) | 1.00 (0.96, 1.00) | 1.00 (0.29, 1.00) | 1.00 (0.96, 1.00) | 0.00 (0.00, NaN) |
| Sample period |  |  |  |  |  |  |  |  |  |  |  |
| April 2020 | 280 | 70 | 200 | 65 | 205 | 0.26 (0.21, 0.32) | 0.90 (0.80, 0.96) | 0.99 (0.96, 1.00) | 0.97 (0.89, 1.00) | 0.97 (0.93, 0.99) | 0.10 (0.05, 0.20) |
| May 2020 | 114 | 3 | 111 | 3 | 111 | 0.03 (0.01, 0.07) | 1.00 (0.29, 1.00) | 1.00 (0.97, 1.00) | 1.00 (0.29, 1.00) | 1.00 (0.97, 1.00) | 0.00 (0.00, NaN) |

